# Signal Mining and Analysis of Adverse Events of Isotretinoin: 20-year real-world pharmacovigilance analysis based on the FAERS database

**DOI:** 10.1101/2025.08.01.25332708

**Authors:** Qian Chen, Mao Zhang, XueDong Kang, LiZhu Han, JinYue Li, Yuan Bian

## Abstract

**Objective:** To identify post-marketing adverse event (AE) signals associated with isotretinoin using real-world data from the U.S. Food and Drug Administration (FDA) AE Reporting System (FAERS), aiming to provide references for clinical safety and further research.

**Methods:** AE reports from the first quarter of 2004 to the third quarter of 2024 were extracted from the FAERS database. Four signal detection methods were employed: the Reporting Odds Ratio (ROR), Proportional Reporting Ratio (PRR), Medicines and Healthcare products Regulatory Agency (MHRA) comprehensive criteria, Bayesian Confidence Propagation Neural Network (BCPNN), and Multi-item Gamma Poisson Shrinker (MGPS).

**Results:** A total of 142,160 AE reports involving isotretinoin were collected, corresponding to 50,519 patients. The four methods identified 469 common AE signals. The top five AEs ranked by descending ROR values were: inflammatory bowel disease (ROR=579.14; 95% CI: 554.95-604.39), gastrointestinal injury (ROR=412.80; 95% CI: 381.18-447.04), fulminant acne (ROR=321.42; 95% CI: 236.39-437.04), ulcerative proctitis (ROR=241.56; 95% CI: 201.70-289.30), and premature epiphyseal closure (ROR=221.22; 95% CI: 172.47-283.74). Among the top 30 AE signals, several conditions, including nasal vestibulitis, anal papilla hypertrophy, neonatal neuroblastoma, diverticular hernia, SAPHO syndrome, somatic delusional disorder, hypersomnia-bulimia syndrome, and hemihypertrophy, were not listed in the drug’s prescribing information. The AE signals involved 25 system organ classes, predominantly psychiatric disorders (75, 15.99%), gastrointestinal disorders (58, 12.37%), and various congenital, familial, and genetic disorders (50, 10.66%). Additionally, strong signals related to pregnancy events were detected, notably unintended pregnancy (ROR=91.39; 95% CI: 86.78-96.26).

**Conclusion:** AE signals associated with isotretinoin involve a broad spectrum of system organ classes. Comprehensive monitoring during clinical use is essential, particularly concerning psychiatric and gastrointestinal disorders. Given the strong signals regarding teratogenicity and pregnancy-related events, strengthening preventive measures for pregnancy risks in patients is recommended.

## 1 Introduction

Isotretinoin is a first-generation, non-aromatic retinoid drug. It significantly improves acne symptoms by inhibiting sebaceous gland sebum secretion, regulating abnormal keratinization of sebaceous gland ducts in hair follicles, exerting anti-inflammatory effects, and preventing scar formation. In 1982, isotretinoin was approved by the FDA for patients aged 12 years and older with severe refractory nodular acne. Clinically, it is also widely used to treat refractory or persistent moderate-to-severe acne, as well as acne prone to scarring or significant psychological distress [0-**Error! Reference source not found**.]. In recent years, isotretinoin has been frequently utilized beyond prescribing indications for psoriasis, cutaneous lupus erythematosus, other skin diseases [**Error! Reference source not found**.,**Error! Reference source not found**.], ichthyosis and other disorders of cornification [**Error! Reference source not found**.], and as maintenance therapy for high-risk neuroblastoma in children [7]. Currently, isotretinoin is commercially available in multiple countries worldwide, and safety concerns regarding its use continue to attract attention. Research indicates a high incidence of adverse reactions involving multiple organ systems, including depression, inflammatory bowel disease (IBD), liver dysfunction, and hypertriglyceridemia [**Error! Reference source not found**.]. However, its association with certain adverse reactions, such as IBD and depression, remains controversial. Therefore, a systematic analysis of large-scale, real-world data is necessary to clarify potential risks. In addition, isotretinoin is teratogenic. Prescribing information from multiple countries strongly warns that women of childbearing age must strictly use contraception while taking isotretinoin. However, the effectiveness of these recommendations requires verification through additional real-world studies. This study will also focus on pregnancy-related events occurring during isotretinoin treatment.

The FAERS system in the United States collects large-scale real-world AE reports through a public voluntary reporting format, characterized by a large sample size and strong timeliness [**Error! Reference source not found**.]. Systematic analysis of the FAERS database enables the identification of potential drug risks, making it a critical tool for drug safety monitoring and pharmacovigilance.

This study conducts signal mining of isotretinoin-related AEs using real-world data from the FAERS database. Consequently, the safety profile of isotretinoin in actual clinical practice can be comprehensively evaluated. The findings of this study aim to provide a scientific basis for clinical drug management, enhance patient medication safety, and assist drug regulatory agencies in identifying, warning, and formulating policies related to potential drug risks.

## 2 Data and Methods

### 2.1 Data sources

This study extracted 83 quarters of data from the FAERS database in the United States, spanning the first quarter of 2004 to the third quarter of 2024. All the data can be accessed at https://fis.fda.gov/extensions/FPD-QDE-FAERS/FPD-QDE-FAERS.html. Definitions for FAERS database fields can be found on the FDA official website. The dataset comprised seven subsets of data, including demographic and administrative information (DEMO), drug/biological information (DRUG), adverse drug reaction information (REAC), patient outcomes information (OUTC), report source information (RPSR), drug therapy start and end dates (THER), and indications for use or diagnosis (INDI).

### 2.2 Data cleaning and standardization

Duplicate, revoked, or deleted reports in the FAERS database required cleaning. First, duplicate reports were removed using the FDA-recommended method. The PRIMARYID, CASEID, and FDA_DT fields from the DEMO table were selected and sorted by CASEID, FDA_DT, and PRIMARYID. For reports with identical CASEID, the record with the latest FDA_DT was retained. For records with identical CASEID and FDA_DT, the report with the highest PRIMARYID was retained. Second, reports identified in the list of deleted reports were removed based on CASEID. Finally, AEs were classified and standardized using preferred terms (PT) and system-organ classification (SOC) from the MedDRA (version 27.1). A comprehensive flowchart outlining the study design is presented in Fig 1.

**Fig 1.**
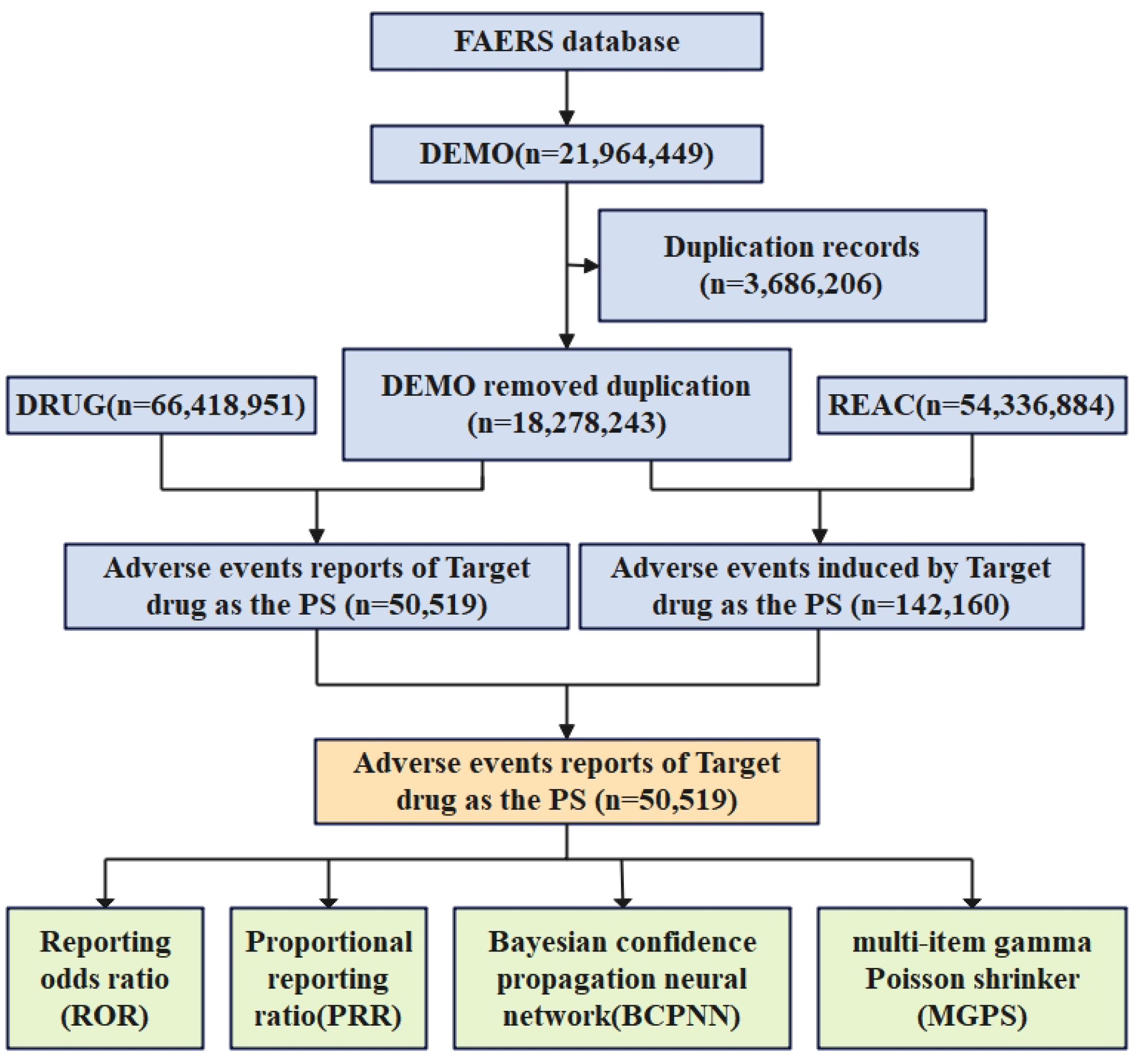
AE analysis process for Isotretinoin using the FAERS database. PS, primary suspect.

### 2.3 Statistical analysis

Data processing and statistical analysis were conducted using Microsoft Excel and SAS 9.4 software. Descriptive analysis was applied to characterize all AE reports related to isotretinoin. Currently, multiple methods are commonly employed to jointly mine AE signals. This study used four signal detection methods to enhance research credibility: ROR, PRR based on the MHRA comprehensive criteria, BCPNN, and MGPS methods [**Error! Reference source not found**.-**Error! Reference source not found**.]. Data were classified and statistically analyzed using a four-grid table (see Table 1). Signals identified as positive by all four detection methods were considered risk signals associated with isotretinoin. The comprehensive criteria for signal determination were: a ≥ 3, lower limit of the ROR 95% confidence interval > 1, PRR ≥ 2 and χ ^2^ ≥ 4, IC-2SD > 0, and EBGM05 > 2. Specific calculation formulas and signal criteria are presented in Table 2.

**Table 1.** Four-grid table for measures of disproportionality.

|  | Target AEs | Non-target AEs | Total |
| --- | --- | --- | --- |
| Target Drug | a | b | a+b |
| Non-target Drug | c | d | c+d |
| Total | a+c | b+d | N=a+b+c+d |

**Table 2.**
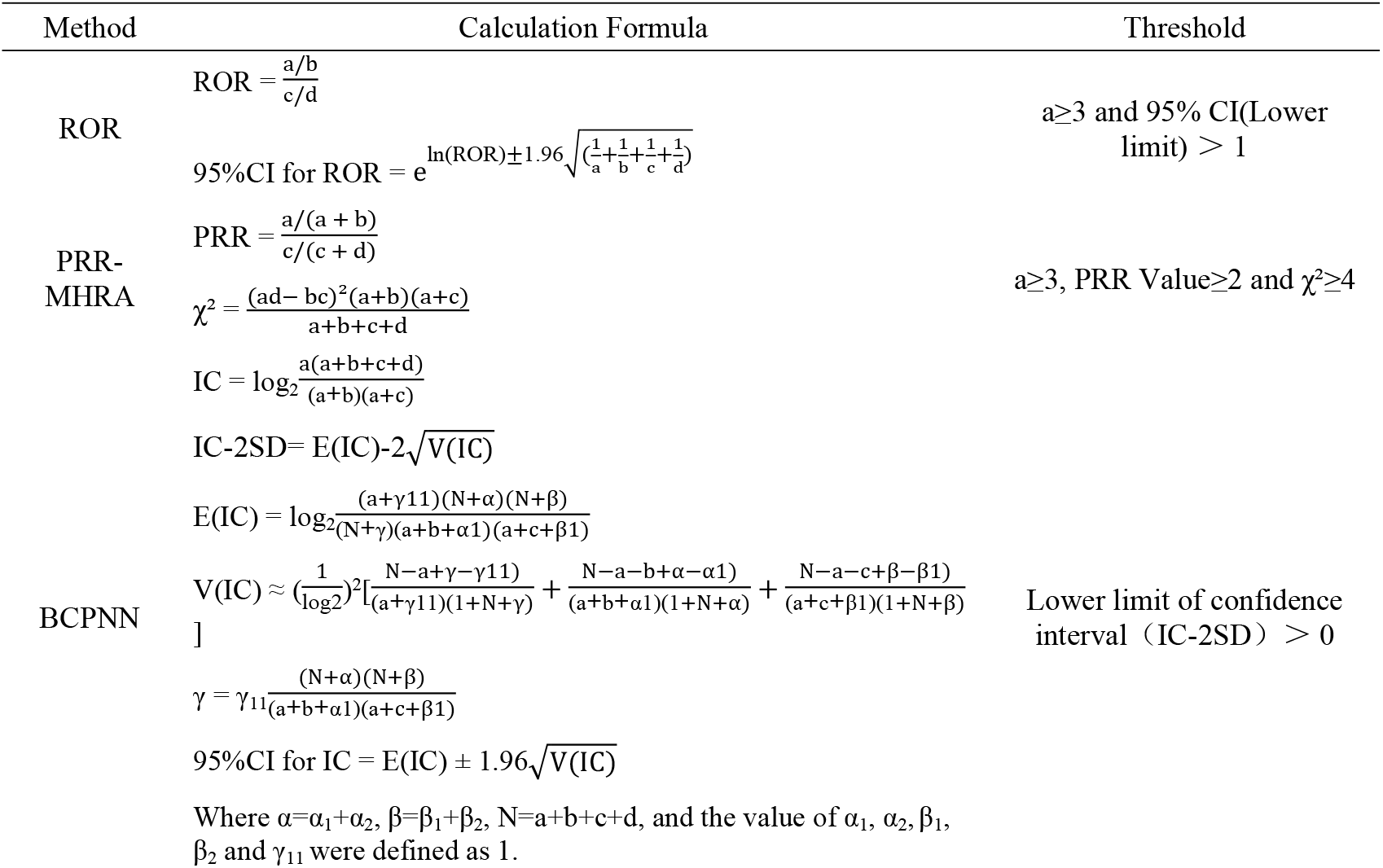
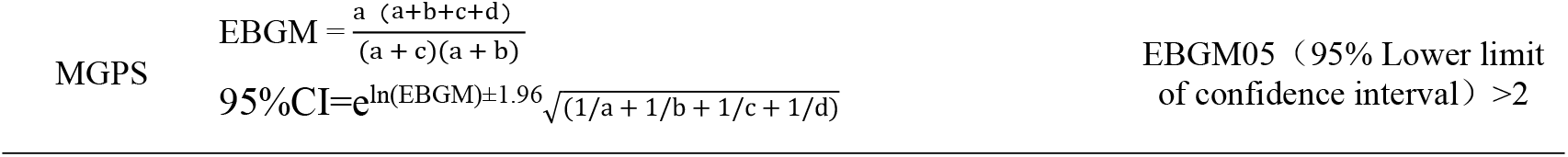
Formulas and threshold values for ROR, PRR-MHRA, BCPNN, and MGPS.

## 3 Results

### 3.1 Basic information and demographic characteristics of AE reports

The number of background patients included in this study was 18,278,243 (with 54,336,884 AEs), while the number of patients administered isotretinoin was 50,519 (with 142,160 AEs). According to statistics, the number of AE reports related to isotretinoin gradually increased each year. Regarding patient gender, females accounted for 55.44%, males for 33.21%, and unknown gender for 11.35%. Patients were predominantly young, with 34.07% aged between 18 and 44 years, and 19.15% younger than 18 years. The United States was the primary reporting country, accounting for 80.14% of reports. Physicians (33.47%) and consumers (29.52%) were the main reporters. Specific details are provided in Table 3.

**Table 3.**
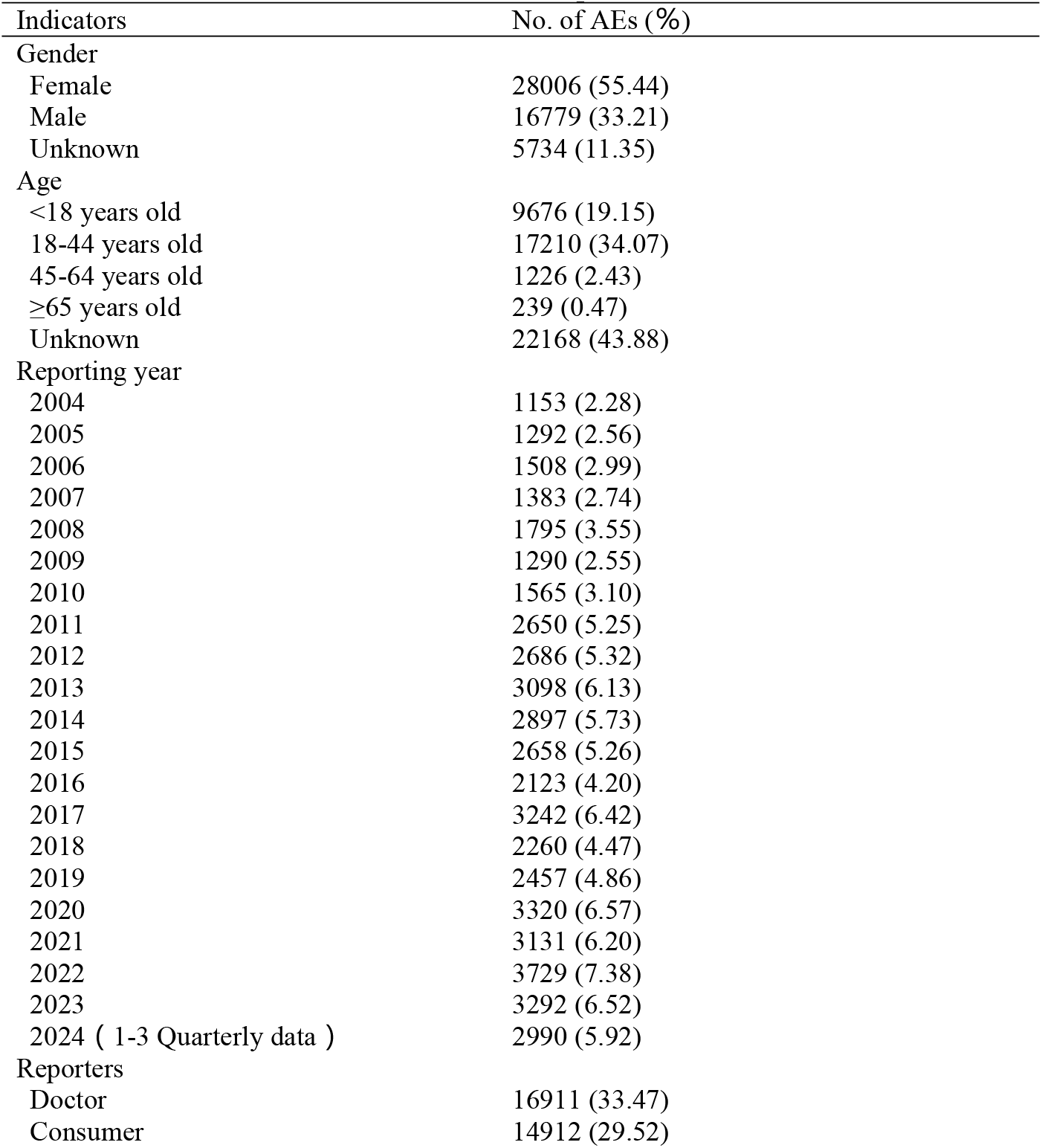
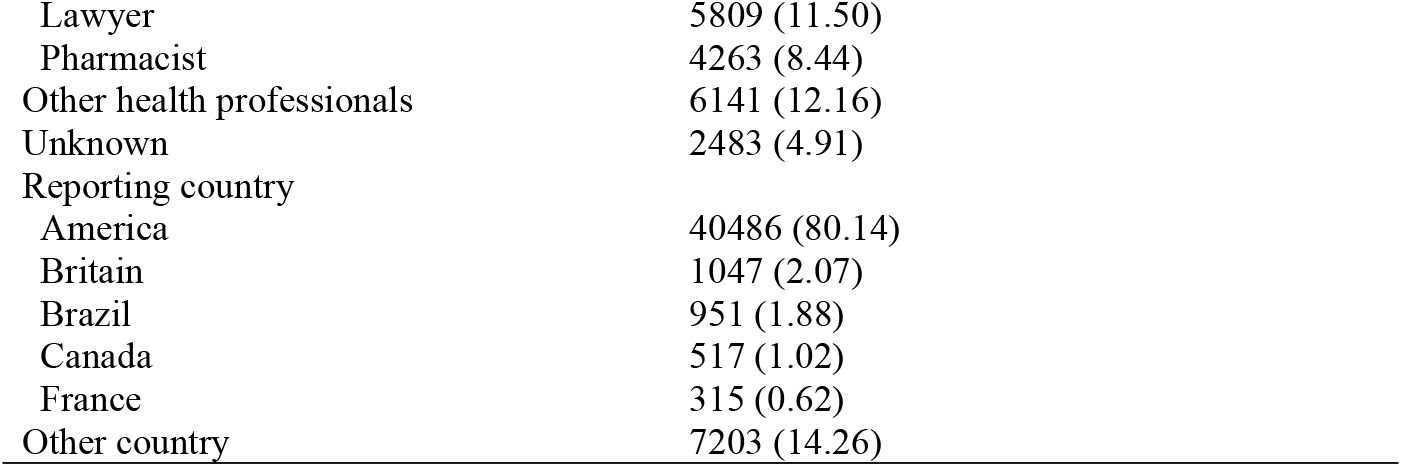
Basic information of AE reports related to isotretinoin.

| Indicators | No. of AEs (%) |
| --- | --- |
| Gender |  |
| Female | 28006 (55.44) |
| Male | 16779 (33.21) |
| Unknown | 5734 (11.35) |
| Age |  |
| <18 years old | 9676 (19.15) |
| 18-44 years old | 17210 (34.07) |
| 45-64 years old | 1226 (2.43) |
| ≥65 years old | 239 (0.47) |
| Unknown | 22168 (43.88) |
| Reporting year |  |
| 2004 | 1153 (2.28) |
| 2005 | 1292 (2.56) |
| 2006 | 1508 (2.99) |
| 2007 | 1383 (2.74) |
| 2008 | 1795 (3.55) |
| 2009 | 1290 (2.55) |
| 2010 | 1565 (3.10) |
| 2011 | 2650 (5.25) |
| 2012 | 2686 (5.32) |
| 2013 | 3098 (6.13) |
| 2014 | 2897 (5.73) |
| 2015 | 2658 (5.26) |
| 2016 | 2123 (4.20) |
| 2017 | 3242 (6.42) |
| 2018 | 2260 (4.47) |
| 2019 | 2457 (4.86) |
| 2020 | 3320 (6.57) |
| 2021 | 3131 (6.20) |
| 2022 | 3729 (7.38) |
| 2023 | 3292 (6.52) |
| 2024 ( 1-3 Quarterly data ) | 2990 (5.92) |
| Reporters |  |
| Doctor | 16911 (33.47) |
| Consumer | 14912 (29.52) |
| Lawyer | 5809 (11.50) |
| Pharmacist | 4263 (8.44) |
| Other health professionals | 6141 (12.16) |
| Unknown | 2483 (4.91) |
| Reporting country |  |
| America | 40486 (80.14) |
| Britain | 1047 (2.07) |
| Brazil | 951 (1.88) |
| Canada | 517 (1.02) |
| France | 315 (0.62) |
| Other country | 7203 (14.26) |

### 3.2 Signal intensity of AEs related to isotretinoin

This study identified 469 positive AE signals related to isotretinoin through the joint application of four methods. The AE signals detected by each method were generally consistent, as shown in Table 4. Furthermore, these results were mostly consistent with AE occurrences documented in isotretinoin prescribing information, confirming the credibility of the analytical method. Ranked in descending order by ROR values, the top ten AEs were IBD, gastrointestinal injury, acne fulminans, ulcerative proctitis, premature epiphyseal closure, nasal vestibulitis, induced complete abortion, selective abortion, xerosis, and epiphyseal injury. Comparing these results with the prescribing information revealed several AEs not previously documented, including nasal vestibulitis, hypertrophic anal papilla, neonatal neuroblastoma, diverticular hernia, SAPHO syndrome, somatic delusional disorder, hypersomnia-bulimia syndrome, and hemihypertrophy.

**Table 4.** The top 30 AE signals related to isotretinoin.

| PTs | No. of AEs | ROR (95% CI) | PRR (Chi-Square) | IC (IC-2SD) | EBGM (EBGM05) |
| --- | --- | --- | --- | --- | --- |
| Inflammatory bowel disease | 5278 | 579.14(554.95-604.39) | 557.68(1191034) | 7.83(7.72) | 227.03(217.54) |
| Gastrointestinal injury | 1260 | 412.80(381.18-447.04) | 409.15(247454) | 7.63(7.32) | 197.86(182.71) |
| Acne fulminans | 75 | 321.42(236.39-437.04) | 321.26(12994.3) | 7.45(5.33) | 174.80(128.56) |
| Proctitis ulcerative | 193 | 241.56(201.70-289.30) | 241.23(28278.8) | 7.21(6.15) | 148.13(123.69) |
| Epiphyses premature fusion | 98 | 221.22(172.47-283.74) | 221.06(13588.9) | 7.13(5.52) | 140.29(109.38) |
| Nasal vestibulitis* | 30 | 219.98(140.35-344.79) | 219.94(4146.20) | 7.13(4.06) | 139.84(89.22) |
| Abortion induced complete | 11 | 209.69(100.47-437.64) | 209.67(1473.84) | 7.08(2.49) | 135.63(64.98) |
| Selective abortion | 59 | 195.66(142.95-267.82) | 195.58(7548.89) | 7.02(4.93) | 129.60(94.69) |
| Xerosis | 339 | 183.75(161.40-209.18) | 183.31(41508.4) | 6.96(6.33) | 124.11(109.02) |
| Epiphyseal injury | 9 | 163.39(74.83-356.76) | 163.38(1016.74) | 6.84(2.16) | 114.67(52.52) |
| Lip dry | 1806 | 143.68(136.09-151.70) | 141.87(184126) | 6.70(6.54) | 103.66(98.19) |
| Pityriasis alba | 3 | 127.08(34.40-469.41) | 127.07(281.44) | 6.58(0.30) | 95.56(25.87) |
| Hypertrophic anal papilla* | 5 | 119.14(43.64-325.21) | 119.13(446.25) | 6.51(1.18) | 91.01(33.34) |
| Neonatal neuroblastoma* | 3 | 114.37(31.47-415.58) | 114.37(259.33) | 6.46(0.31) | 88.21(24.27) |
| Anomaly of middle ear congenital | 4 | 108.92(35.85-330.92) | 108.92(332.67) | 6.41(0.80) | 84.94(27.96) |
| Diverticular hernia* | 3 | 103.97(29.01-372.69) | 103.97(240.38) | 6.36(0.32) | 81.91(22.85) |
| Childhood depression | 3 | 103.97(29.01-372.69) | 103.97(240.38) | 6.36(0.32) | 81.91(22.85) |
| Anotia | 18 | 100.92(60.03-169.68) | 100.91(1407.92) | 6.32(3.22) | 80.00(47.58) |
| Cheilosis | 9 | 98.04(47.12-203.95) | 98.03(687.55) | 6.29(2.15) | 78.18(37.58) |
| Acne conglobata | 13 | 95.31(51.90-175.03) | 95.31(970.49) | 6.26(2.73) | 76.44(41.63) |
| Congenital cerebellar<br>agenesis | 8 | 95.31(43.92-206.84) | 95.31(597.23) | 6.26(1.96) | 76.44(35.23) |
| Unintended<br>pregnancy | 1788 | 91.39(86.78-96.26) | 90.26(127634) | 6.19(6.06) | 73.17(69.48) |
| Dermatitis papillaris<br>capillitii | 3 | 87.98(25.07-308.74) | 87.97(209.59) | 6.16(0.33) | 71.67(20.42) |
| Abortion induced | 1600 | 87.93(83.26-92.86) | 86.95(110710) | 6.15(6.01) | 70.99(67.22) |
| Drug exposure before<br>pregnancy | 168 | 81.58(69.05-96.37) | 81.48(11003.5) | 6.07(5.35) | 67.31(56.98) |
| SAPHO syndrome* | 36 | 81.23(56.68-116.40) | 81.21(2351.09) | 6.07(4.07) | 67.12(46.84) |
| Delusional disorder,<br>somatic type * | 3 | 76.25(22.07-263.38) | 76.24(185.64) | 5.99(0.34) | 63.70(18.44) |
| Hypersomnia-bulimia<br>syndrome* | 4 | 72.62(24.93-211.55) | 72.61(237.31) | 5.93(0.82) | 61.16(20.99) |
| Chapped lips | 580 | 67.64(61.92,73.90) | 67.37(32230.3) | 5.84(5.58) | 57.40(52.54) |
| Hemihypertrophy* | 3 | 63.54(18.72,215.71) | 63.54(158.28) | 5.77(0.35) | 54.60(16.08) |

Note: AE signals are sorted in descending order of ROR, with "\*" indicating AEs not recorded in isotretinoin prescribing information.

**Table 5.**
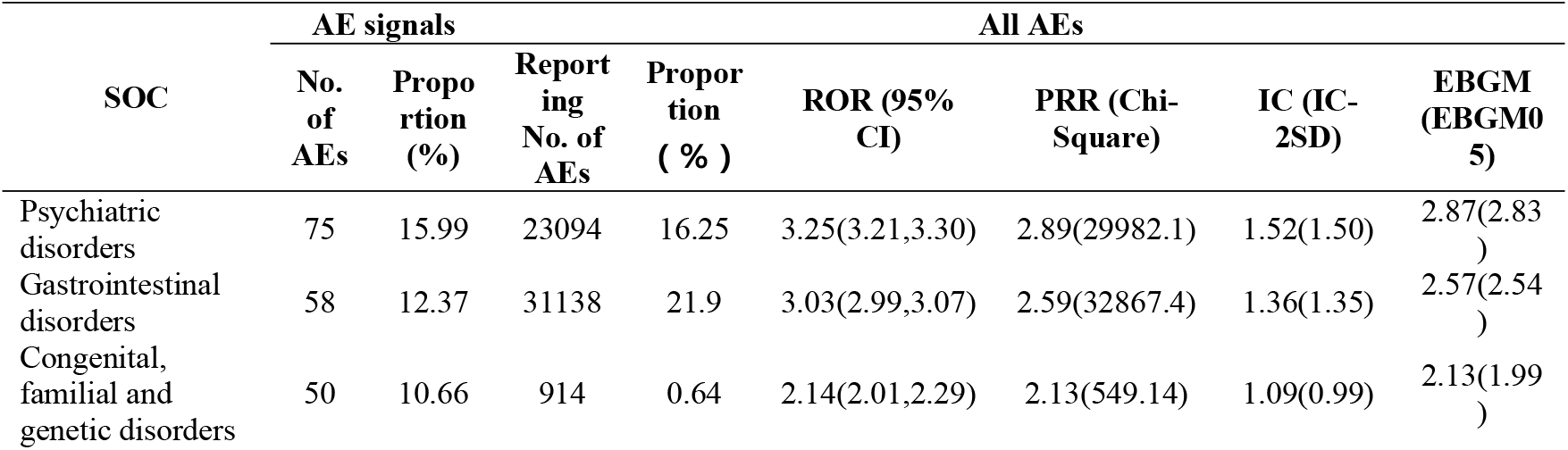
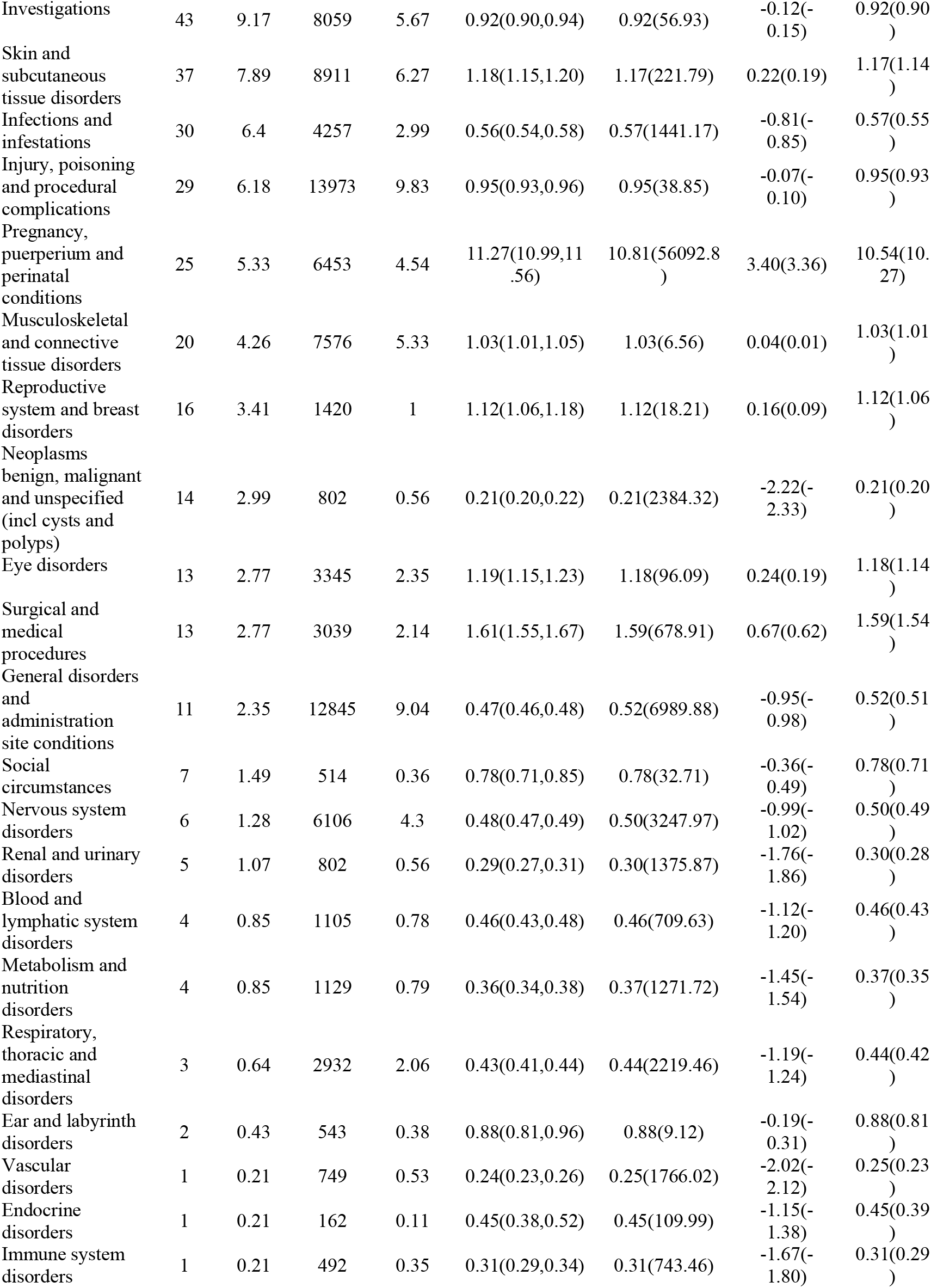
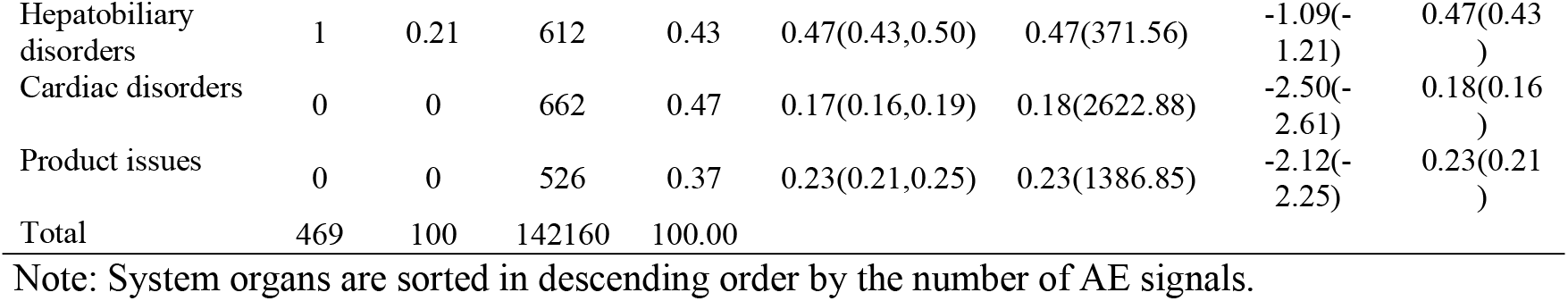
Distribution of SOC involved in isotretinoin-related AEs and cumulative signal intensity for each SOC.

### 3.3 Classification of system organs involved in AE signals related to isotretinoin

AE reports involved 27 organs classes within the SOC system. The highest number of AE reports was in Gastrointestinal disorders (31,138 cases, 21.90%), followed by Psychiatric disorders (23,094 cases, 16.25%) and Injury, poisoning and procedural complications (13,973 cases, 9.83%).

Positive AE signals involved 25 organ classes in the SOC system. Ranked by the number of signals, the top three were: Psychiatric disorders (75, 15.99%), Gastrointestinal disorders (58, 12.37%), and Congenital, familial, and genetic disorders (50, 10.66%).

Four methods were employed to detect cumulative signals across various SOC organ classes. Positive cumulative signals were identified in three organ classes. Ranked in descending order of ROR, these were: Pregnancy, puerperium, and perinatal conditions (ROR 11.27, 95% CI lower limit 10.99), Psychiatric disorders (ROR 3.25, 95% CI lower limit 3.21), and Gastrointestinal disorders (ROR 3.03, 95% CI lower limit 2.99).

### 3.4 Onset Time of AEs

After excluding reports with inaccurate, missing, or unknown onset times, a total of 12,575 AEs associated with isotretinoin provided valid onset-time data. The results indicated a median AE onset time of 72 days (interquartile range [IQR]: 26–171 days), with more than half of the cases occurring within the first three months of treatment (0–30 days: n = 3,742, 29.76%; 31–60 days: n = 1,872, 14.89%; 61–90 days: n = 1,436, 11.42%). Moreover, the incidence of AEs after six months of treatment was considerable, showing an increasing trend and accounting for 23.98% of the total cases (181–360 days: n = 1,107, 8.80%; >360 days: n = 1,909, 15.18%). The timeline of these events is illustrated in Fig 2, and the cumulative incidence curve of AEs is presented in Fig 3.

**Fig 2.**
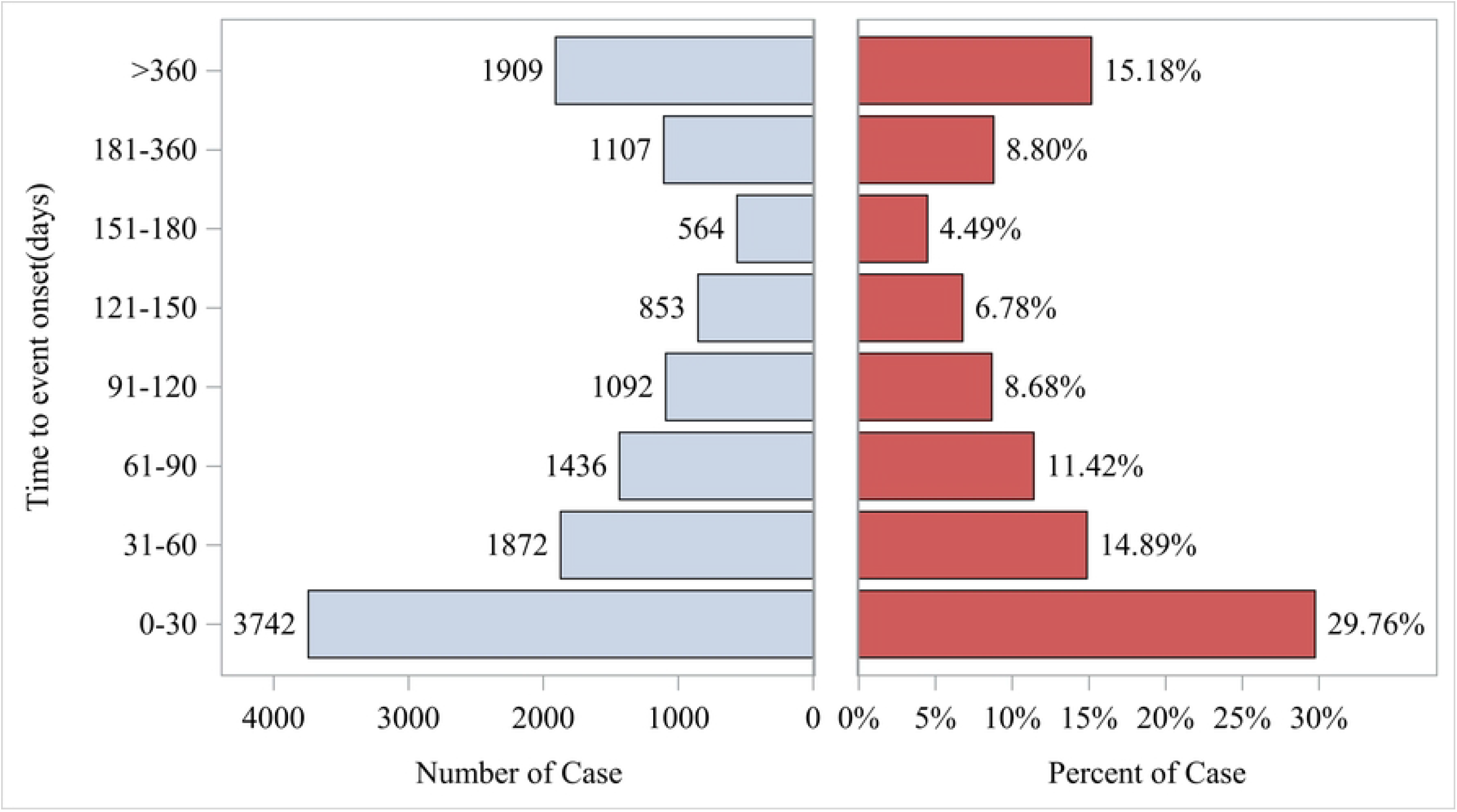
Time to event report distribution of AE reports.

**Fig 3.**
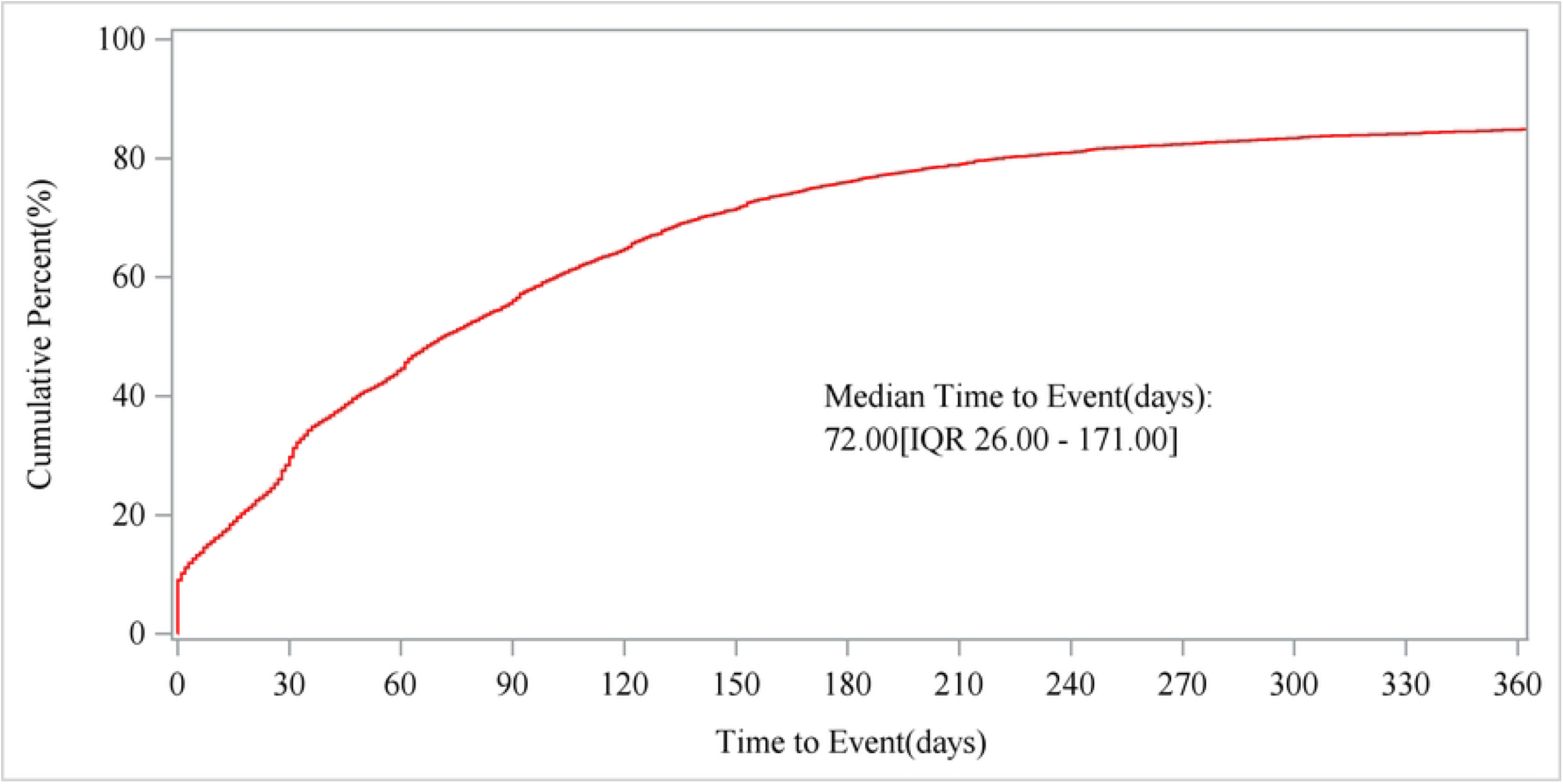
Cumulative incidence of AEs.

## 4 Discussion

### 4.1 Basic characteristics of AE reports related to isotretinoin

Patients included in the isotretinoin-related AE reports of this study were predominantly young adults and minors, with females comprising the majority. This might be associated with the higher incidence of acne in post-adolescent women compared to men, and the significantly higher proportion of women seeking medical treatment. Studies have indicated that during adolescence, acne incidence is roughly equal between genders. However, prevalence is higher among females post-adolescence [**Error! Reference source not found**.,**Error! Reference source not found**.]. Due to factors including appearance-related anxiety, sociocultural pressures, and physiological characteristics [**Error! Reference source not found**.], women are more inclined to seek professional treatment [**Error! Reference source not found**.], such as prescription medications or long-term therapies. Men, by contrast, typically seek medical attention only in severe cases.

### 4.2 Characteristics of isotretinoin-related AEs involving SOC

This study identified AE signals involving 25 SOC systems, suggesting isotretinoin-related AEs potentially affect multiple organ systems. Previous research has demonstrated that isotretinoin treatment impacts nearly all human body systems, resulting in various AEs [**Error! Reference source not found**.]. This conclusion aligns closely with our study findings. During clinical use, comprehensive monitoring and timely interventions are essential. SOC systems with high signal numbers and AE reports primarily included psychiatric disorders, gastrointestinal disorders, congenital, familial, and genetic disorders, investigations, and skin and subcutaneous tissue disorders. These results are largely consistent with prior reports [**Error! Reference source not found**.] and with isotretinoin prescribing information, supporting the feasibility and reliability of this research methodology.

AE signals and reports for psychiatric and gastrointestinal disorders ranked highest, greatly exceeding other SOCs. These findings indicate that AEs in these categories are most frequent clinically. Prominent PT signals within psychiatric disorders (sorted by descending ROR) included childhood depression, somatic delusional disorder, hypersomnia-bulimia syndrome, purging behaviors, and emotional changes related to eating disorders. Prominent PT signals within gastrointestinal disorders (sorted by descending ROR) included IBD, ulcerative proctitis, cheilosis, hypertrophic anal papilla, and diverticular hernia.

Analysis of cumulative signals across SOC organ systems revealed that pregnancy, puerperium, and perinatal conditions exhibited the strongest signals identified by all four detection methods. A total of 6,453 events were reported in this category, among which 4,641 (71.92%) were pregnancy-related events, presenting robust positive signals. These results highlight the necessity of careful monitoring for pregnancy-related events during isotretinoin treatment in clinical practice.

### 4.3 Onset time of isotretinoin-associated adverse events

The findings of this study indicated that the median onset time of adverse events in patients treated with isotretinoin was 72 days, with the majority of cases occurring within the first three months after initiating therapy. Furthermore, the incidence of AEs exhibited an upward trend after six months of treatment. Thus, in clinical practice, careful monitoring for AEs is particularly important within the initial three months of treatment as well as after six months of continued isotretinoin use to ensure patient safety.

### 4.4 AE signals of key concern

#### 4.4.1 Isotretinoin and AE signals related to psychiatric disorders

Currently, the correlation between isotretinoin and psychiatric disorders remains controversial. Studies suggest that isotretinoin may increase anxiety and depression risks by reducing serotonin levels, increasing neuronal apoptosis, regulating gene expression, affecting neural plasticity, or activating the HPA axis [**Error! Reference source not found**.]. However, other research indicates that isotretinoin is mainly prescribed for moderate to severe acne, a condition associated with a higher prevalence of psychiatric disorders [**Error! Reference source not found**.], especially in adolescents and young adults. Patients frequently experience depression, anxiety, and suicidal tendencies due to altered physical appearance and social anxiety [**Error! Reference source not found**.]. Thus, patients prescribed isotretinoin inherently have a higher risk of psychiatric disorders. When investigating psychiatric side effects of isotretinoin, the influence of acne itself must be comprehensively considered [**Error! Reference source not found**.]. Recent large-scale cohort studies and meta-analyses have not definitively confirmed a causal relationship [**Error! Reference source not found**.].

Although existing evidence is insufficient to establish causality between isotretinoin and psychiatric disorders, the association cannot be completely excluded. Monitoring of psychiatric AEs is currently recommended in both domestic and international prescribing information for isotretinoin. This study identified strong signals for psychiatric AEs based on extensive real-world data from isotretinoin users, warranting serious attention. Clinically, mental health screening before isotretinoin therapy is recommended. Psychological status should be regularly monitored throughout treatment. Personalized management strategies should be adopted for high-risk patients. Psychological support and regular follow-up should be provided during therapy to mitigate psychiatric risks. Education on medication for patients and families must be strengthened. Patients displaying psychiatric symptoms should seek prompt medical attention. Multidisciplinary management approaches should be enhanced, including collaboration with psychiatric or psychological specialists if necessary.

#### 4.4.2 Isotretinoin and IBD

This study found that IBD ranked first in signal intensity and second in reported events, demonstrating exceptionally strong signal strength. Some studies indicate isotretinoin may induce IBD by disrupting intestinal barrier function and activating the immune system. However, large-scale population studies over the past decade, recent meta-analyses, and clinical guidelines suggest that IBD onset strongly correlates with genetic, environmental, and immune factors. These confounders commonly exist in study populations, and thus a direct causal relationship between isotretinoin and IBD remains unproven [**Error! Reference source not found**.,**Error! Reference source not found**.-**Error! Reference source not found**.].

Given these conclusions, the current study further analyzed possible reasons for the strong correlation between isotretinoin and IBD. After 2009, multiple lawsuits against isotretinoin manufacturers occurred in the United States, significantly increasing reported IBD cases between 2009 and 2012. Prior studies indicate that a large proportion of FAERS reports linking isotretinoin to IBD were submitted by law firms, suggesting potential legally driven overreporting [**Error! Reference source not found**.,**Error! Reference source not found**.].

Further analysis of the raw data showed that 5,278 IBD reports existed, among which 4,483 (84.94%) were submitted by lawyers. Analyzing reporting years revealed that 4,526 cases (85.75%) were reported between 2009 and 2014. Since 2018, fewer than 10 reports per year have been submitted, again highlighting potential legally driven overreporting after 2009. Considering these factors, the strong positive IBD signal found in this study likely reflects significant reporting bias. Therefore, larger prospective studies are needed to further investigate the relationship between isotretinoin and IBD. Nevertheless, close monitoring of gastrointestinal symptoms during isotretinoin use remains advisable, particularly for patients with a family history of IBD or those diagnosed with IBD.

#### 4.4.3 Teratogenicity of isotretinoin

Isotretinoin can cross the placental barrier and is classified as Grade X by the FDA regarding pregnancy safety. Clinical studies have shown that oral administration of isotretinoin increases the risk of severe congenital malformations, spontaneous abortion, and premature birth [**Error! Reference source not found**.]. This study found isotretinoin ranked third among positive signals involving congenital, familial, and genetic disorders, mainly affecting the ears and labyrinth, central nervous system, cardiovascular system, musculoskeletal system, and eyes. This finding aligns with the drug prescribing information and existing literature [**Error! Reference source not found**.,**Error! Reference source not found**.]. The top ten identified congenital abnormalities included congenital middle ear abnormalities, anorectal malformations, congenital cerebellar hypoplasia, congenital optic nerve abnormalities, hemihypertrophy, congenital inner ear abnormalities, cerebellar dysplasia, anophthalmia, congenital ear malformations, and follicular keratosis. Special attention should be given to hemihypertrophy and follicular keratosis, which have strong positive signals but are not mentioned in isotretinoin prescribing information. Current studies suggest potential associations between isotretinoin exposure and partial developmental asymmetry, unilateral abnormal growth, skin dysplasia, and keratinization disorders [**Error! Reference source not found**.,**Error! Reference source not found**.], though definitive correlations remain unconfirmed. Further research should explore these relationships.

#### 4.4.4 Isotretinoin and pregnancy events

This study reported 6,453 AE occurrences related to pregnancy, puerperium, and perinatal conditions, with 4,641 (71.92%) specifically categorized as pregnancy events. Among these, unexpected pregnancy displayed strong positive signals, ranking within the top 30 AE signals. These findings highlight the necessity of enhanced management of pregnancy events in clinical practice. Isotretinoin carries a high risk of severe birth defects and is contraindicated in pregnant women or those who may soon become pregnant. According to prescribing information in some countries, such as China, women of childbearing age or their spouses must use effective contraception within three months before isotretinoin use, during treatment, and for three months after cessation. The US FDA guidelines require contraception for at least one month before starting medication, during treatment, and for three months after discontinuation. Pregnancy prevention programs, such as the iPLEDGE program in the United States, have been implemented internationally to reduce isotretinoin-associated fetal malformation risks by ensuring strict contraception before and after administration [**Error! Reference source not found**.,**Error! Reference source not found**.].

Despite these programs, 3.3%–6.5% of isotretinoin users still experience pregnancy during treatment [**Error! Reference source not found**.]. Key reasons include insufficient patient compliance with contraception, inadequate education regarding pregnancy prevention, and improper contraceptive method selection. Research suggests some patients fail to fully understand isotretinoin’s severe teratogenic risks before treatment. Combined with poor contraceptive compliance, this leads to a high incidence of unintended pregnancies [**Error! Reference source not found**.].

Based on these findings, clinical doctors or pharmacists must remind patients to strictly use contraception during and after isotretinoin treatment. Patients should prioritize highly effective contraceptive methods or multiple contraceptive strategies. Pregnancy monitoring should be strengthened, and enhanced patient education is essential for improving compliance. Government regulatory agencies should further strengthen and improve pregnancy prevention measures to ensure the safe use of isotretinoin.

## 5 Research Innovations and Limitations

### 5.1 Research Innovations

#### 5.1.1 Methodological Innovation

We pioneered the systematic integration of four internationally recognized pharmacovigilance algorithms (ROR, PRR, BCPNN, MGPS). This integrated approach was applied to a comprehensive analysis of 142,160 adverse event reports associated with isotretinoin spanning two decades (2004– 2024) from the FAERS database. This significantly enhanced the robustness of signal detection and the identification capability for rare events.

#### 5.1.2 Expansion of the Risk Profile

We established the most comprehensive adverse event profile for isotretinoin to date, identifying 469 consolidated safety signals. Notably, 8 previously unlisted signals (including nasal vestibulitis, anal papilla hypertrophy, and neonatal neuroblastoma) offer novel evidence for updating global safety information in prescribing documents.

#### 5.1.3 Novel Risk Management Findings

Unintended pregnancy (ROR=91.39) emerged as the strongest pregnancy-related signal. This finding provides critical empirical evidence for optimizing global Pregnancy Prevention Programs (e.g., iPLEDGE) and highlights potential vulnerabilities in current risk management protocols.

### 5.2 Research Limitations

Although this study analyzed a large amount of real-world data, certain limitations remain. (1) The data analyzed were primarily reported from European and American countries, potentially influenced by racial and regional differences. (2) Due to the spontaneous reporting nature of the database, some information might be misreported or underreported, potentially affecting research results. (3) Detected AE signals indicate only a statistical correlation between the drug and AE, and their clinical significance requires further verification through clinical trials.

## 6 Conclusions

In summary, this study utilized the FAERS database in the United States and employed SAS 9.4 software with four detection methods to identify AE signals associated with isotretinoin in the real world. Findings indicated that AEs were primarily concentrated in psychiatric and gastrointestinal disorders, aligning with the prescribing information for isotretinoin. In clinical practice, monitoring and follow-up should be enhanced. The study also identified AE signals not documented in the prescribing information, including nasal vestibulitis, hypertrophic anal papilla, neonatal neuroblastoma, diverticular hernia, SAPHO syndrome, somatic delusional disorder, hypersomnia-bulimia syndrome, and hemihypertrophy, which require special attention. Additionally, this study detected extremely high-risk signals related to severe birth defects and pregnancy events. Therefore, further efforts are necessary to strengthen medication education for patients and preventive management measures concerning pregnancy events to ensure medication safety. Due to inherent difficulties in quantitative analysis based on real-world data, these findings still require verification through large-scale prospective studies.

## Data Availability

All the source data can be accessed at https://fis.fda.gov/extensions/FPD-QDE-FAERS/FPD-QDE-FAERS.html. Definitions for FAERS database fields can be found on the FDA official website. All relevant data are within the manuscript and its Supporting Information files.

## Author Contributions

Conceptualization, Q.C. and X.K.; methodology, Q.C. and M.Z.; data curation, J.L. and L.H.; writing—original draft preparation, Q.C.; writing—review and editing, Y.B.; All authors have read and agreed to the published version of the manuscript.

## Institutional Review Board Statement

Ethical approval was not required as the study involved analysis of publicly available anonymized data that posed no risk to participants. According to local regulations and institutional requirements, this human subjects research did not require ethical approval. In compliance with national regulations and institutional policies, written informed consent from participants or their legal guardians/next of kin was not required for this study.

## Informed Consent Statement

Informed consent was not required given the exclusive use of anonymized, non-identifiable data.

## Data Availability Statement

The data that support the findings of this study are available from the corresponding author upon reasonable request.

## Data Availability

All raw data required to replicate the study results are included in the manuscript and Supporting Information files. The data used in the analysis are publicly available via the FDA FAERS database, as stated in the Methods section: https://fis.fda.gov/extensions/FPD-QDE-FAERS/FPD-QDE-FAERS.html.

